# Trans-ancestry polygenic models for the prediction of LDL blood levels: An analysis of the UK Biobank and Taiwan Biobank

**DOI:** 10.1101/2023.08.03.23293320

**Authors:** Emadeldin Hassanin, Ko-Han Lee, Tzung-Chien Hsieh, Rana Aldisi, Yi-Lun Lee, Dheeraj Bobbili, Peter Krawitz, Patrick May, Chien-Yu Chen, Carlo Maj

## Abstract

**Background:** Polygenic risk scores (PRSs) are proposed for use in clinical and research settings for risk stratification. PRS predictions often show bias toward the population of available genome-wide association studies, which is typically of European ancestry. This study aims to assess the performance differences of ancestry-specific PRS and test the implementation of multi-ancestry PRS to enhance the generalizability of low-density lipoprotein (LDL) cholesterol predictions in the East Asian population

**Methods:** We computed ancestry-specific and multi-ancestry PRS for LDL using data from the global lipid consortium while accounting for population-specific linkage disequilibrium patterns using PRS-CSx method. We first conducted an ancestry-wide analysis using the UK Biobank dataset (n=423,596) and then applied the same models to the Taiwan Biobank dataset (TWB, n=68,978). PRS performances were based on linear regression with adjustment for age, sex, and principal components. PRS strata were considered to assess the extent to which a PRS categorization can stratify individuals for LDL cholesterol levels in East Asian samples.

**Results:** Population-specific PRS better predicted LDL levels within the target population but multi-ancestry PRS were more generalizable. In the TWB dataset, covariate-adjusted R^2^ values were 9.3% for ancestry-specific PRS, 6.7% for multi-ancestry PRS, and 4.5% for European-specific PRS. Similar trends (8.6%, 7.8%, 6.2%) were observed in the smaller East Asian population of the UK Biobank (n=1,480). Consistent with the R^2^ values, PRS stratification in East Asians (TWB) effectively captured a heterogenous variability in LDL blood cholesterol levels across PRS strata. The mean difference in LDL levels between the lowest and highest East Asian-specific PRS (EAS_PRS) deciles was 0.82, compared to 0.59 for European-specific PRS (EUR_PRS) and 0.76 for multi-ancestry PRS. Notably, the mean LDL values in the top decile of multi-ancestry PRS were comparable to those of EAS_PRS (3.543 vs. 3.541, *P*=0.86).

**Conclusions:** Our analysis of the PRS prediction model for LDL cholesterol further supports the issue of PRS generalizability across populations. Our targeted analysis of the East Asian (EAS) population revealed that integrating non-European genotyping data, accounting for population-specific linkage disequilibrium, and considering meta-analyses of non-European-based GWAS alongside powerful European-based GWAS can enhance the generalizability of LDL PRS.

## Background

Blood lipid levels are a significant, modifiable, and heritable risk factors for coronary artery disease (CAD), including low-density lipoprotein (LDL-C)^1^. Previous studies have demonstrated that lipid levels have a moderate-to-high heritability variations, ranging from 20 to 60%^2^. Numerous common variants have been discovered in recent genome-wide association studies (GWASs) associated with LDL, as well as many other traits^3^. However, the majority of these variants are weakly associated individually with a given trait or disease and have limited predictive power. The cumulative effects of several common variants have been suggested to contribute significantly to the risk stratification for clinical utility. Methods have been developed for analyzing data from these large-scale studies and detecting genetic variants and phenotype associations, i.e., one such method is the polygenic risk score (PRS). Several studies have evaluated the association between PRS and the risk of various conditions^4^, including lipid traits^5^, CAD^6^, cancer^7,8^, diabetes^9^, and neurodevelopmental disorders.

One of the major issues concerning the translational use of PRS is the strong dependency on population specificity. In fact, the performance of PRS can be significantly influenced by the linkage-disequlibrium (LD) across variants and allele frequencies that are specific to different populations^10^. As a consequence, PRS has been mostly limited to European ancestry cohorts for which larger reference GWAS are available^11^. In addition to LD and allele frequencies also gene-environment interactions might also be responsible for the different genetic susceptibilities toward a trait. For instance the genetic liability of lipid levels is less understood in East Asian ancestry populations^11^. Since individuals with East Asian ancestry account for more than a fifth of the global population, understanding genetic variation in East Asians is crucial to improve risk characterization and preventive interventions^12^.

In the last few years the availability of large population-based cohorts and cross-ancestry GWAS enabled also the development of novel computational algorithms to improve the generalizability of PRS^13,14^. A multi-ancestry, GWAS meta-analysis of lipid levels was conducted by the Global Lipid Genetics Consortium including 350,000 people of non-European ancestry, 150,000 East Asian individuals, and approximately 1.65 million people^5^. The study also demonstrated that our understanding of the genetic component associated with lipid levels is significantly improved by increasing diversity rather than including additional European ancestry individuals.

In this study, we derived ancestry-specific and cross-ancestry PRS to predict serum LDL level by first considering all populations and then focusing on East Asian individuals. Specifically, we derived six LDL-PRSs: four ancestry-specific PRS (East Asian, South Asian, European, African) and two multi-ancestry PRSs (East Asian with European meta-analysis, and the four ancestry meta-analysis). The six PRSs were tested among nine population groups estimated from the UK Biobank (UKB, n=423,596). We focused on the East Asian ancestry group from the UKB and validated the PRS with participants from the Taiwan Biobank (TWB, n=68,978). We then tested the associations between PRS and LDL cholesterol changes among East Asian individuals in both biobanks.

## Methods

### Study subjects

The analysis was performed using genetic and phenotypic data of the UKB and TWB. UKB is a population-based cohort study, with over 500,000 individuals aged 40 to 69 years at the time of recruitment^15^. We excluded outliers with high genotype missing rates, putative sex chromosome aneuploidy, and discordant reported sex vs genotypic sex^16^. We randomly excluded one from each pair of related individuals if the genetic relationship was closer than the second degree, defined as kinship coefficient > 0.0884 as calculated by the UKB. We applied a previous approach to divide UKB individuals into nine ancestry groups by projecting data onto the PCA space of 1000 Genomes Project^11^.

TWB is a Taiwanese-based cohort study, with 68,978 individuals aged from 30 to 75 across 750k SNPs^17^. For more overlapping SNPs with PRS models, we imputed the TWB cohort. First, we filtered out SNPs based on the missing rate and Hardy-Weinberg equilibrium. Then, we imputed the genotype with a reference based on the whole genome sequencing data of 1,496 Taiwanese individuals. In total, we obtained 15 million SNPs for 69k Taiwanese individuals as our external validation set.

### UK Biobank ancestry grouping

We assigned the samples to different countries using PC-projection as demonstrated in a previous study^11^. In this previous study, the authors explored different methods to classify individuals into ancestry groups using principal component analysis (PCA) of genome-wide genotype data. They find that Euclidean distances in the PCA space are proportional to genetic differences between populations and recommend using this distance measure. They suggest using all principal components to capture population structure, as using only two or four is insufficient for distinguishing certain populations. They apply PCA-based distance to infer ancestry in datasets and propose two solutions: projecting PCs to reference populations or using internal data. They demonstrate that these solutions are effective for inferring ancestry and grouping genetically similar individuals. Here, we used this approach to define the nine ancestry groups based on UK Biobank data and birth country information, with some groups including individuals from neighboring countries (namely, East Asian: China; European: United Kingdom, Italy, and Poland; African: Nigeria, and Caribbean; South Asian: India; Middle East: Iran, and Ashkenazi Jewish). Additionally, we defined East Asian subpopulations by projecting samples in the 1000 Genomes Project PC space considering the five East Asian subpopulations as references.

### Construction of multi-ancestry polygenic score

To evaluate the potential of PRS to predict increased LDL cholesterol levels in East Asian ancestry. We used the latest Global Lipid Genetics Consortium GWAS that was conducted in different populations to derive an ancestry-specific or multi-ancestry LDL PRS^5^. We considered the summary statistics that did not include UK Biobank samples. Six PRSs were created: one for each ancestry (East Asian, South Asian, European, African), and two meta-analyses using multi-ancestry GWAS (one using East Asian and European ancestry, and the other using the four ancestries). PRSs weights were conducted using PRS-CSx^13^ (accounting for population-specific allele frequencies and LD patterns) and the 1000 Genomes Project as a reference panel that matched the ancestry of each discovery GWAS. The PRS-CSx method incorporates summary statistics from different GWAS and links the genetic effects across populations using a continuous shrinkage prior that is shared between them. This approach allows for more precise estimation of effect sizes by using information from the summary statistics and taking advantage of the variation in linkage disequilibrium across the discovery samples. By jointly modeling these multi-ancestry summary statistics, PRS-CSx might be able to better capture the underlying genetic effects and produce more accurate predictions. We developed the multi-ancestry PRS using the “--meta” option provided by the software. We tested each of the six PRSs in the nine population groups from the UKB. We then evaluated the six PRSs among the East Asian cohort of the TWB. We compared the PRS performance between individuals in TWB and two East Asian sub-populations from the UKB (namely Han Chinese South [CHS] and Kinh in Ho Chi Minh City, Vietnam [KHV]) from the 1000 Genomes Project. Notebly, that most of TWB individuals are clustered with the CHS group^18^. We excluded the other three East Asian subpopulations due to sample size limitations.

### Assessment of PRS accuracy

We assessed the prediction accuracy of the six PRSs in the nine estimated populations from the UKB and Taiwanese population from the TWB. We standardized PRSs to a mean of 0 and standard deviation of 1. Two models were used: (1) the full model which included PRS with sex, age, age^2^ and the first four genetic principal components as covariates (formula: LDL ∼ PRS + sex + age + age^2^ + PC1 + PC2 + PC3 + PC4), and (2) the reference model which only accounted for covariates (formula: LDL ∼ sex + age + age^2^ + PC1 + PC2 + PC3 + PC4). Linear regression was performed, and incremental R^2^ was calculated following previous study^19^ as the difference between the adjusted R^2^ of the full model (including PRS as an additional predictor) and the reference model. Mean LDL values across deciles of EAS_PRS, EUR_PRS, and multi-ancestry PRS were computed in all individuals of TWB to evaluate the range of phenotypic variability cover for these PRS.

## Results

### Study populations

In the UK Biobank, the estimated ethnic groups of the United Kingdom (UK) and China had significantly different study participant characteristics (Table 1). In comparison to people in the United Kingdom (UK), Chinese participants had lower LDL concentrations (mean, SD: 3.42 mmol/L, 0.77), lower TC levels (mean, SD: 5.54 mmol/L, 1.03), and similar HDL levels (mean, SD: 1.46 mmol/L, 0.38). They were also younger (mean age, SD: 52.3, 7.71). The Chinese participants had a lower percentage of men than the UK (38.8% vs. 45.9%). Participants from China had a significantly lower body mass index (BMI) (mean, SD: 24.07 kg/m2, 3.4) than UK participants (*p*-value <2.2 × 10^−16^).

**Table 1.** Study participant characteristics stratified by estimated ethnicity in UK Biobank. For continuous variables, p-values from the Welch t-statistic tests are reported, while for categorical and binary variables, p-values from Pearson’s Chi-squared tests are reported. HC, hypercholesterolemia; HDL, high-density lipoprotein cholesterol; LDL, low-density lipoprotein cholesterol; SD, standard deviation.

|  | Participants, N | Males, N (%) | Age, Mean (SD) | HC Cases, N (%) | HC Controls, N | BMI, Mean (SD) | LDL, Mean (SD) | HDL, Mean (SD) | TC, Mean (SD) |
| --- | --- | --- | --- | --- | --- | --- | --- | --- | --- |
| **UK Biobank** |  |  |  |  |  |  |  |  |  |
| **United Kingdom** | 423596 | 194259 (45.9) | 56.81 (8.02) | 110166 (26.01) | 313430 (73.99) | 27.4 (4.76) | 3.57 (0.87) | 1.45 (0.38) | 5.71 (1.14) |
| **Italy** | 6451 | 2882 (44.7) | 54.5 (8.41) | 1624 (25.17) | 4827 (74.83) | 27.35 (4.94) | 3.56 (0.86) | 1.45 (0.38) | 5.68 (1.12) |
| **India** | 6303 | 3413 (54.1) | 53.42 (8.41) | 1135 (18.01) | 5168 (81.99) | 27.42 (4.5) | 3.35 (0.85) | 1.25 (0.32) | 5.31 (1.12) |
| **Poland** | 4095 | 1544 (37.7) | 54.4 (7.53) | 1088 (26.57) | 3007 (73.43) | 27.39 (4.96) | 3.59 (0.85) | 1.49 (0.4) | 5.76 (1.13) |
| **Nigeria** | 3802 | 1744 (45.9) | 51.95 (8.14) | 551 (14.49) | 3251 (85.51) | 29.82 (5.31) | 3.21 (0.84) | 1.43 (0.35) | 5.17 (1.09) |
| **Caribbean** | 2492 | 898 (36) | 52.52 (8.13) | 396 (15.89) | 2096 (84.11) | 29.49 (5.56) | 3.28 (0.83) | 1.47 (0.38) | 5.29 (1.09) |
| **Ashkenazi** | 2359 | 1067 (45.2) | 58.09 (7.1) | 613 (25.99) | 1746 (74.01) | 27.13 (4.54) | 3.55 (0.9) | 1.44 (0.39) | 5.68 (1.2) |
| **China** | 1480 | 545 (36.8) | 52.33 (7.71) | 263 (17.77) | 1217 (82.23) | 24.07 (3.4) | 3.42 (0.77) | 1.46 (0.38) | 5.54 (1.03) |
| **Iran** | 1145 | 680 (59.4) | 51.99 (7.98) | 234 (20.44) | 911 (79.56) | 27.98 (4.55) | 3.43 (0.86) | 1.28 (0.33) | 5.4 (1.11) |
| **P-Value: United Kingdom/China** |  | <1.1 x 10<sup>-16</sup> | <2.2 x 10<sup>-16</sup> | <2.9 x 10<sup>-16</sup> | <2.9 x 10<sup>-16</sup> | <2.2 x 10<sup>-16</sup> | <1.8 x 10<sup>-13</sup> | 0.53 | <1.6 x 10<sup>-10</sup> |
| **Taiwan Biobank** |  |  |  |  |  |  |  |  |  |
| **Taiwan** | 68,978 | 21,495 (31.2) | 51.0 (10.9) | 8,196 (13.5) | 60,782 (86.5) | 24.25 (3.8) | 3.16 (0.82) | 1.43 (0.35) | 5.12 (0.93) |

In the TWB, the percentage of men is 31.2% which is lower than the percentage of Chinese participants in the UK Biobank, while the age distribution (mean, SD: 51.0, 10.9) is similar. In addition, TWB individuals had lower levels of lipid traits, including LDL (mean, SD: 3.16 mmol/L, 0.82), HDL (mean, SD: 1.43 mmol/L, 0.35), and TC (mean, SD: 5.12 mmol/L, 0.93), but higher BMI (mean, SD: 24.25 kg/m^2^, 3.80).

### Evaluation of the PRS in the nine-estimated populations from the UK Biobank

We assessed the performance of ancestry-specific PRS for the LDL levels across the nine estimated populations in the UKB (Figure 1). As expected, the LDL PRS derived from European GWAS (EUR_PRS) was associated with the best performance in different European populations (namely, United Kingdom, Poland, and Italy) and in Middle East populations (namely, Ashkenazi Jews and Iranians). Similarly, the LDL PRS derived from African GWAS (AFR_PRS) showed the best performance in the population of African origin (Nigeria and Caribbean). The LDL PRS derived from the East Asian GWAS (EAS_PRS) was the best performing in the Chinese population. Surprisingly, when we tested EUR_PRS and PRS derived from the South Asian GWAS (SAS_PRS) in the India participants, EUR_PRS performed better than SAS_PRS.

**Figure 1.**
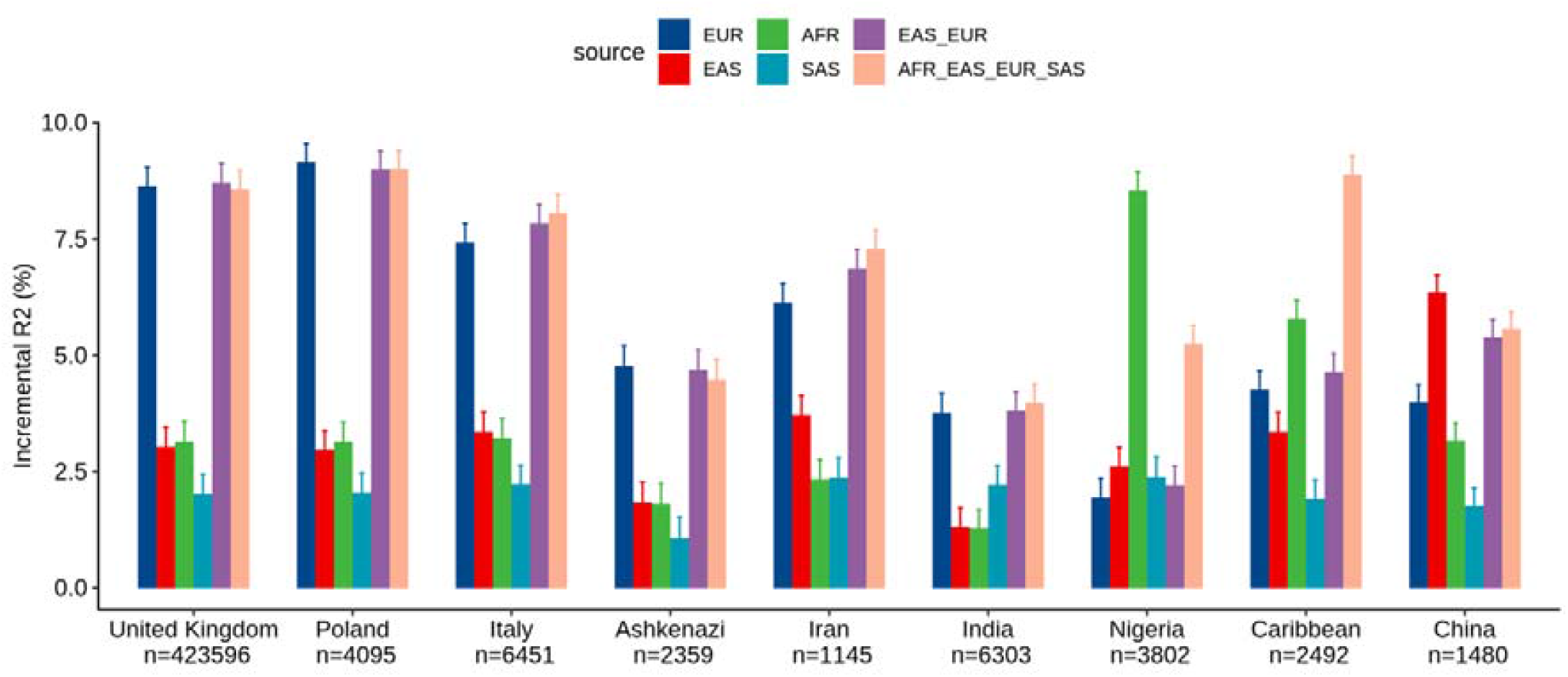
Performance of ancestry-specific and multi-ancestry PRS models for LDL across nine sub-population in the UKB.

Concerning the multi-ancestry PRS, we tested a PRS derived from a meta-analysis of European and East Asian GWASs (EUR_EAS_PRS), and a global PRS derived from a meta analysis of the four ancestries (EUR_EAS_SAS_AFR_PRS). The multi-ancestry PRS showed comparable prediction to ancestry-specific PRS and seems to be more generalizable across populations, specifically for European, Middle East, and SAS populations. For instance, for the United Kingdom population, the adjusted R^2^% using EUR_PRS (8.62%) was similar to that using EUR_EAS_SAS_AFR_PRS (8.56%). For the AFR and EAS populations, ancestry-specific PRS performed better than multi-ancestry PRS. For instance, for the Chinese population, the adjusted R^2^% using EAS_PRS (6.35%) was higher than that using EUR_EAS_SAS_AFR_PRS (5.55%).

### Evaluation of the PRS in the Taiwan biobank

Within the TWB, we evaluated the different ancestry-specific and multi-ancestry PRSs for the LDL levels (Figure 2). Similar to our findings in the UKB China participants, the EAS_PRS (adjusted R^2^%=9.3%) also demonstrated better performance than EUR_PRS (adjusted R^2^%=4.5%) in the TWB individuals and had even a better performance compared to multi-ancestry PRS (adjusted R^2^%=6.7%). We also compared the performance of PRS between TWB individuals and the East Asian sub-populations from the UKB. We found that EAS_PRS has a comparable performance specially between populations from TWB (adjusted R^2^%=6.5%) and CHS (adjusted R^2^=6.1%) from UKB.

**Figure 2.**
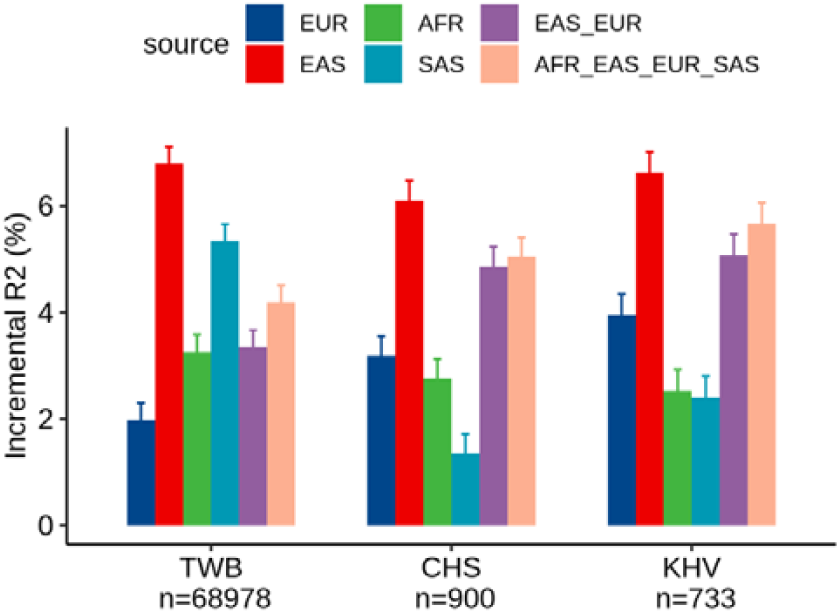
Performance of ancestry-specific and multi-ancestry PRS models for LDL on East Asian populations.

### Association between different PRS strata and LDL values

We analyzed the mean of LDL levels in individuals from TWB based on their EAS_PRS, EUR_PRS, and multi-ancestry PRS deciles. We compared the difference in mean LDL levels between the lowest and highest deciles of EAS_PRS, EUR_PRS, and Multi PRS. Our findings showed that in East Asians, EAS_PRS explained a wider range of phenotypic variability compared to EUR_PRS. Specifically, the difference in mean LDL levels between the lowest and highest EAS_PRS deciles was 0.82, while for EUR_PRS it was 0.59 (Figure 3). The mean difference in LDL levels between the lowest and highest multi-ancestry PRS deciles was 0.76. However, the mean LDL levels in the highest deciles in both EAS_PRS and multi-ancestry PRS were the same (LDL mean (mmol/L) = 3.54, *P*=0.86).

**Figure 3.**
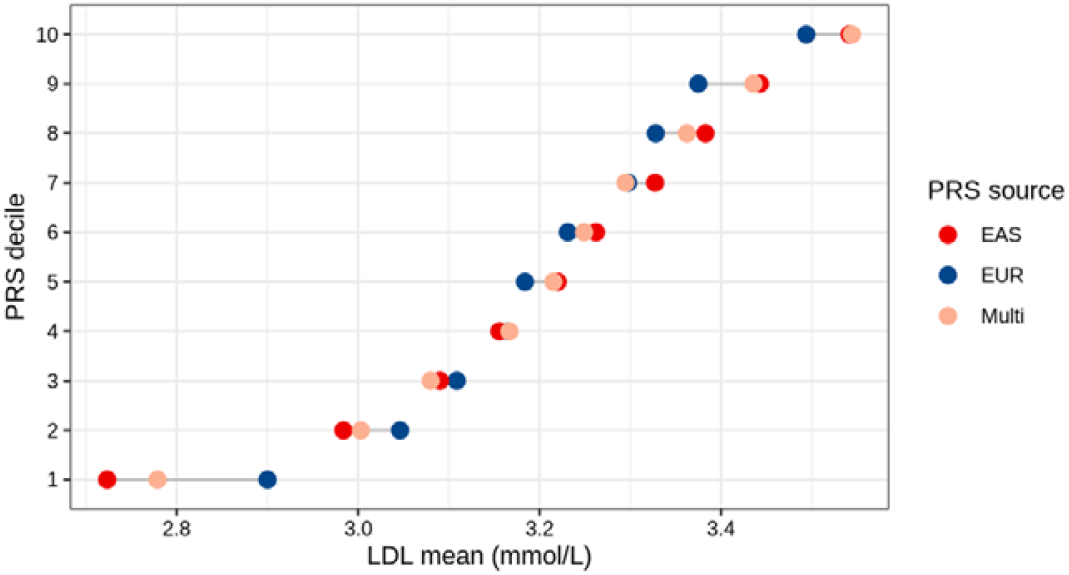
LDL mean values across deciles of EAS, EUR and Multi PRSs of East Asians (TWB) PRS.

## Discussion

In this study aimed at predicting LDL in two EAS populations (from UKB and TWB) using the latest Global Lipids Genetics Consortium GWAS. Our findings indicate that: 1) ancestry-specific PRS yield better performance in predicting LDL levels, and 2) multi-ancestry PRSs together with computational approaches integrating population specific LD-pattern can be used to enhance the generalizability of PRSs. In particular, the multi-ancestry PRSs showed that even relatively small proportions of non-European samples can significantly improve prediction in non-EUR populations. Our work emphasizes the importance of conducting global GWAS that include diverse populations to enhance the generalizability of PRSs, even when the availability of diverse population samples is limited.

Our study further suggest that statistical genetics approaches can be used to take advantage of the already available global GWAS data, even when the number of non-European samples is limited. One example, the latest Global Lipids Genetics Consortium GWAS includes individuals across five genetic ancestry groups: admixed African or African (6.0% of the sample), East Asian (8.9%), European (79.8%), Hispanic (2.9%), and South Asian (2.5%)^5^. Recently published Bayesian PRS approaches demonstrated an improvement in the accuracy of PRSs in non-European populations by utilizing common genetic effects across ancestries^13,14^.

Another recent study, the authors conducted a benchmarking analysis to compare several PRS methods for multi-ancestry analysis in the UKBB dataset, which included lipid traits and EAS data using also global lipid consortium GWAS data^20^. The findings of this study provided insights on the use of statistical methods to improve prediction performance in non-Europeans.

The applicability of the findings on the portability of PRS from multi-ancestry meta-analyses to other traits needs to be taken into account, considering multiple factors^21^. These factors include the heritability of the trait^22^, genetic correlation^23^, causal variants allele frequencies^24^, gene-environment interactions^25^, and the inclusion of multi-ancestry populations in GWAS^26,27^. In a recent study, they estimated the cross-ancestry genetic correlation for cholesterol and observed a significant genetic heterogeneity between ancestries for total and LDL cholesterol^22^. While many traits exhibit a significant shared genetic correlation across ancestries, indicating potential transferability of multi-ancestry PRS^28^, some traits have specific genetic variations that are more commonly found in particular ancestral groups^29,30^. To ensure the effective use of PRS in diverse populations, it is crucial to conduct comprehensive investigations considering these factors and include a representative range of ancestries in future GWAS studies ^31^. Moreover, a recent study emphasizes the necessity of moving away from discrete genetic ancestry clusters and embracing the continuum of genetic ancestries when analyzing and interpreting PGS^10^. By accounting for individual variation and considering the diverse genetic backgrounds within populations, more accurate PGS assessments can be achieved.

By leveraging the available diverse GWAS data, we can improve the generalizability of PRSs, and ultimately enhance our ability to predict complex disease risk across diverse populations. As such, our study provides valuable insights into the development and implementation of PRSs for predicting lipid traits in East Asian populations, and highlights the need for continued efforts to increase diversity in genetic research while also working on bioinformatics approaches to meta-analyze the association signal across different populations.

## Conclusion

In our study we evaluated the performance of ancestry-specific and multi-ancestry PRSs for LDL, in various populations including East Asians from the UK Biobank and Taiwan Biobank. The findings corroborated that ancestry specific PRSs performed better than out of target population PRSs in the respective ancestry. In particular, the EAS_PRS had better performance in East Asian populations, while the EUR_PRS showed better performance in European and Middle East populations. The multi-ancestry PRS analysis showed that even a small proportion of non-European samples can significantly improve prediction in non-EUR populations. These findings provide valuable insights into the development of PRSs for diverse populations and the potential clinical applications of PRSs. On the one hand, our analysis suggests that incorporating cross-ancestry GWAS data and utilizing optimized computational algorithms to account for population-specific LD-patterns can improve the generalizability of PRS. On the other hand, these results further emphasize the necessity of enhancing genetic diversity in GWAS studies and establishing large-scale population-based cohorts to more accurately model the genetic liability of multifactorial traits, such as LDL cholesterol.

## Data Availability

Access to genome-wide genotyping data, and phenotypic data from the UK and Taiwan Biobank can be obtained through a successful project application process. Detailed information about the application process can be found at (http://www.ukbiobank.ac.uk/about-biobank-uk/) and (https://taiwanview.twbiobank.org.tw/data_appl). Certian restrictions apply to the availability of these data, as they were used under license for the current study.

## List of abbreviations

PRS: Polygenic risk scores
UKB: UK
TWB: Taiwan Biobank
TC: total cholesterol
LDL-C: low-density lipoprotein cholesterol
GWAS: genome-wide association studies
CHS: Han Chinese South
KHV: Kinh in Ho Chi Minh City, Vietnam
EUR: European
EAS: East Asian SAS: South Asian
AFR: African
PC: Principal Component

## Declarations

### Ethics approval and consent to participate

This study made use of anonymized data from UK Biobank and Taiwan Biobank. Written informed consent was provided by all individuals. The research ethics committee for the UK Biobank and Taiwan Biobank gave its approval to the protocol and the permission. Our investigation was carried out in accordance with approved UK Biobank data application number 52446 and Taiwan Biobank data application number TWBR10411-03. We hereby confirm that, all methods were carried out in accordance with relevant guidelines and regulations.

### Consent for publication

Not Applicable

### Availability of data and code

Access to genome-wide genotyping data, and phenotypic data from the UK and Taiwan Biobank can be obtained through a successful project application process. Detailed information about the application process can be found at (http://www.ukbiobank.ac.uk/about-biobank-uk/) and (https://taiwanview.twbiobank.org.tw/data_appl). Certain restrictions apply to the availability of these data, as they were used under license for the current study. The codes related to the statistical analysis for this study have been deposited on GitLab at the following location (<u>doi:10.17881/8wqn-x712</u>).

### Authors’ contributions

EH, KL, YL and CM performed the statistical analysis and the bioinformatics. EH, KL, CC and CM conceived and designed the study. EH, KL, CC and CM drafted the initial manuscript. EH, KL, TH, RA, YL, DRB, PK, PM, CC and CM performed the critical expert revision. CC and CM supervised the study. All authors read and approved the final manuscript.

## Acknowledgements

UK Biobank analyses were conducted via application 52446 using a protocol approved by the Partners HealthCare Institutional Review Board and Taiwan Biobank using data application number TWBR10411-03.

